# Time series analysis of daily data of COVID-19 reported cases in Japan from January 2020 to February 2023

**DOI:** 10.1101/2023.04.19.23288796

**Authors:** Ayako Sumi

**Affiliations:** Division of Physics, Department of Liberal Arts and Sciences, Center for Medical Education, Sapporo Medical University, Sapporo, Hokkaido 060-8556, Japan

## Abstract

This study investigatbed temporal variational structures of the COVID-19 pandemic in Japan using a time series analysis incorporating maximum entropy method (MEM) spectral analysis, which produces power spectral densities (PSDs). This method was applied to daily data of COVID-19 cases in Japan from January 2020 to February 2023. The analyses confirmed that the PSDs for data in both the pre- and post-Tokyo Olympics periods show exponential characteristics, which are universally observed in PSDs for time series generated from nonlinear dynamical systems, including the so-called susceptible/exposed/infectious/recovered (SEIR) model, well-established as a mathematical model of temporal variational structures of infectious disease outbreaks. The magnitude of the gradient of exponential PSD for the pre-Olympics period was smaller than that of the post-Olympics period, because of the relatively high complex variations of the data in the pre-Olympics period caused by a deterministic, nonlinear dynamical system and/or undeterministic noise. A 3-dimensional spectral array obtained by segment time series analysis indicates that temporal changes in the periodic structures of the COVID-19 data are already observable before the commencement of the Tokyo Olympics and immediately after the introduction of mass and workplace vaccination programs. Lessons from theoretical studies for measles control programs may be applicable to COVID-19.

## Introduction

Since December 2019, a novel coronavirus designated as Severe Acute Respiratory Syndrome Coronavirus 2 (SARS-CoV-2) has rapidly spread around the world, affecting millions of people worldwide; its impact continues today. Waves of cases of this novel coronavirus disease, also known as COVID-19, still occur recursively, although these waves may well be prevented, and possibly eradicated, in the future. Considerable effort to prevent and eradicate COVID-19 has been expended through COVID-19 surveillance, vaccinations, and theoretical and experimental research [1-4]. Among these efforts, attempts to elucidate the mechanism of the COVID-19 pandemic have been of great interest. Recently, researchers have tried to interpret the behavior of the pandemic in terms of deterministic chaos [5-7]. Sapkota et al., reported that the hosting of the Tokyo Olympic Games in Japan between 23 July and 8 August 2021 affected the mechanism of the COVID-19 pandemic in the country [8]. Therefore, the temporal variational structures of the data before and after the Olympic Games may differ, and examining this point is significant from the standpoint of predicting the COVID-19 pandemic. However, typical approaches cannot fully elucidate the temporal variational structures of the patterns of the pandemic. This is because the data lengths of reported cases of COVID-19 are very short: in Japan, the data are collected daily, and the number of data points was slightly above 1000 points by February 2023. Thus, a superior and powerful method of time series analysis to elucidate the temporal variational structure of time series of even short data length is required. In previous publications [9-11], we proposed a method that enables us to analyze the time series of COVID-19 cases. In the present study, this method was applied to examining the temporal variational structures of daily time series data of reported COVID-19 cases for the entire country of Japan. Quantitative elucidations of the COVID-19 pandemic are of great significance in epidemiology, and will contribute to the development of surveillance of the disease.

## Methods

### Data

The present study analyzed daily data of reported COVID-19 cases for the entire country of Japan from 16 January 2020 to 21 February 2023 (1133 data points). During this period, a total 32,851,731 cases of COVID-19 were reported in Japan. The data used in the present study were obtained from the Japan Ministry of Health, Labour, and Welfare COVID-19 Data [12]. The data are indicated in S1 Dataset.

### Time series analysis

We used a time series analysis consisting of maximum entropy method (MEM) spectral analysis in the frequency domain and least squares method (LSM) in the time domain [9-11,13,14]. The MEM is considered to have a high degree of resolution of spectral estimates compared with other analysis methods of infectious disease surveillance data such as the fast Fourier transform algorithm and autoregressive methods, which require time series of long data lengths [15, 16]; therefore, an MEM spectral analysis allows us to precisely determine short data sequences, such as the infectious disease surveillance data used in this study [9-11,13-15].

### MEM spectral analysis

We assumed that the time series data x(t) (where t is time) were composed of systematic and fluctuating parts [17]:

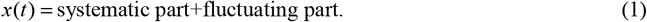

To investigate temporal patterns of x(t) in the monthly time series data, we performed MEM spectral analysis [9-11,13,14]. MEM spectral analysis produces a power spectral density (PSD), from which we obtain the power representing the amount of amplitude of *x*(*t*) at each frequency (note the reciprocal relationship between the scales of frequency and period). The MEM-PSD, *P*(*f*) (where *f* represents frequency), for the time series with equal sampling interval Δ*t*, can be expressed by

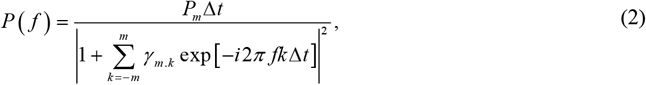

where the value of *P*_*m*_ is the output power of a prediction-error filter of order *m* and *γ*_*m, k*_ is the corresponding filter order. The value of the MEM-estimated period of the *n*th peak component *T*_*n*_ (=1/*f*_*n*_; where *f*_*n*_ is the frequency of the *n*th peak component) can be determined by the positions of the peaks in the MEM-PSD.

### Least squares method

The validity of the MEM spectral analysis results was confirmed by calculating the least squares fitting (LSF) curve pertaining to the original time series data *x*(*t*) with MEM-estimated periods. The formula used to generate the LSF curve for *X*(*t*) was as follows:

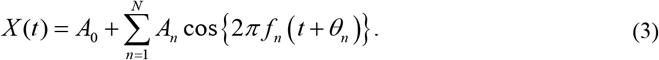

The above formula is calculated using the LSM for *x*(*t*) with unknown parameters *f*_*n*_, *A*_0_ and *A*_n_ (*n* = 1, 2, 3, …, *N*), where *f*_*n*_ (= 1/*T*_*n*_; *T*_*n*_ is the period) is the frequency of the *n*-th component; *A*_0_ is a constant that indicates the average value of the time series data; *A*_*n*_ is the amplitude of the *n*-th component; *θ*_*n*_ is the phase of the *n*-th component; and *N* is the total number of components. The reproducibility level of *x*(*t*) by the optimum LSF curve was evaluated via a Spearman’s rank correlation (ρ) analysis performed using SPSS (Statistical Package for the Social Sciences) version 17.0J software (SPSS, Tokyo, Japan). A *p* value of ≤ 0.05 was considered statistically significant.

## Results

### Temporal Variations in COVID-19 Reported Cases Data

Daily data of reported cases of COVID-19 from 16 January 2020 to 21 February 2023 in Japan are plotted in Fig 1a. A closer view of the data from January 2020 to June 2021 is illustrated in Fig 1b, which shows four large waves observed at intervals of about four to five months, peaking in April and July 2020, and January and May 2021. Subsequently, Fig 1a shows four large waves at longer intervals than before, approximately five to six months, with peaks in August 2021, February and August 2022, and January 2023.

**Fig 1.**
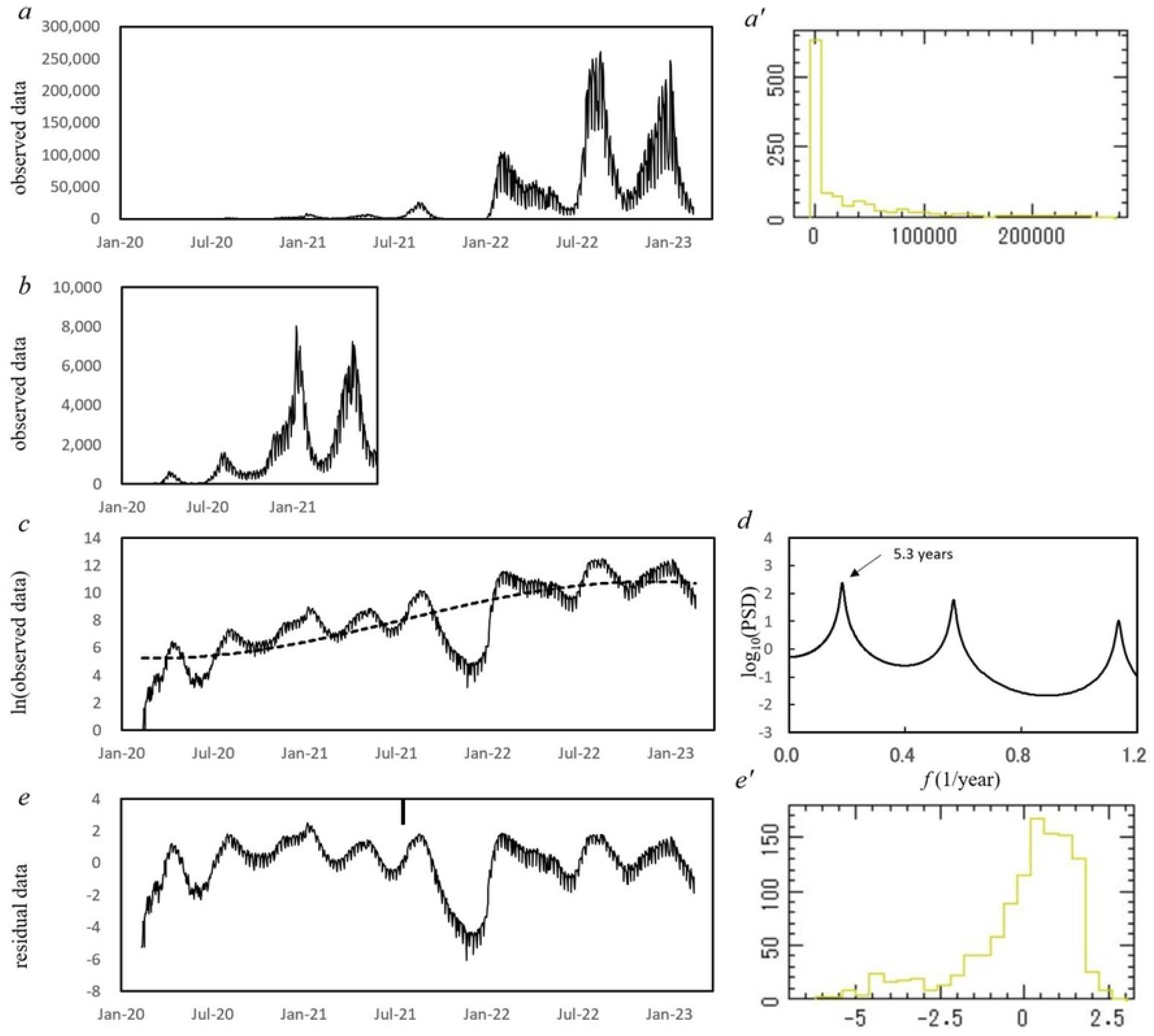
Daily data of COVID-19 reported cases of Japan from 16 January 2020 to 21 February 2023. (a) The original data. (a’) Histogram of the original data. (b) Enlargement of the original data from 16 January 2020 to 30 June 2023. (c) The logarithm-transformed data of the original data (thin line) and its optimum LSF curve (bold line). (d) MEM-PSD of the residual data in the low-frequency range (*f ≤* 1.2). (e) The residual data obtained by subtracting the LSF curve from the log-data. (e’) Histogram of the residual data. Small vertical line in *e* indicates the boundary of phase I (pre-Tokyo Olympic Games, 11 February 2020–22 July 2021) and phase II (post-Tokyo Olympic Games, 23 July 2021–21 February 2023).

### Preparing the data for analysis

We take the reported cases data *x*(*t*) (*t*: time) to represent discrete ones at *t* = *kΔt* (*k* = 1, 2, 3, …*N*) where *Δt* is the time interval and *N* the length of the time series (*Δt* = 1-day and *N* = 1133, in the present study). Fig 1a′ gives the frequency histogram for *x*(*t*) (Fig 1a). This histogram is apart from the normal distribution required for conventional spectral analysis. Then, we introduced the logarithm transformation of *x*(*t*) (Fig 1a). Because the data from 16 January to 10 February 2020 include 17 zero values and are not available for the logarithm transformation, the 15 cases that were reported during this period are ignored and the starting point of the data was re-set to 11 February 2020. As a result, the present study uses the data from 11 February 2020 to 21 February 2023. For the data in this period, the logarithm-transformed data is shown in Fig 1c, where the spikiness of the reported cases observed in *x*(*t*) has been reduced, and a long-term decreasing trend is observed.

To remove the long-term trend of the log-data shown in Fig 1c, PSD, *P*(*f*) (*f*: frequency), for the log-data was calculated, and the PSD (*f*≤1.2) is displayed in Fig 1d (unit of *f*: 1/year). Therein, the longest period can be observed as the prominent peak at the position of 5.3-year period. With this 5.3-year period, we modeled the long-term trend in the COVID-19 pandemic by calculating the LSF curve (Equation (3)) for the entire log-data (Fig 1c). The LSF curve obtained (Fig 1c) expresses the long-term trend of the log-data well.

We removed the LSF curve from the log-data (Fig 1c), and the residual time series data were obtained, as shown in Fig 1e. The frequency histogram for this residual data is shown in Fig 1e′ and approximates to the normal distribution required for conventional spectral analysis. Normality of distribution was assessed using the χ^2^ fitting test, and the null hypothesis was not rejected (*P* = 0.76).

### Power spectral density of the time series data

To investigate the effect of hosting the Tokyo Olympic Games on the temporal variational structures of the COVID-19 pandemic in Japan, the residual data (Fig 1e) were divided into two ranges (phases I and II) in accordance with the starting and ending points of the Tokyo Olympic Games (23 July 2021 and 8 August 2021, respectively): pre-Olympic Games (11 February 2020 – 22 July 2021) for phase I and post-Olympic Games (23 July 2021 – 21 February 2023) for phase II. MEM-PSDs for the residual data of phases I and II were calculated. The semi-log plots of the PSD are shown in Fig 2a and 2b for phases I and II, respectively.

**Fig 2.**
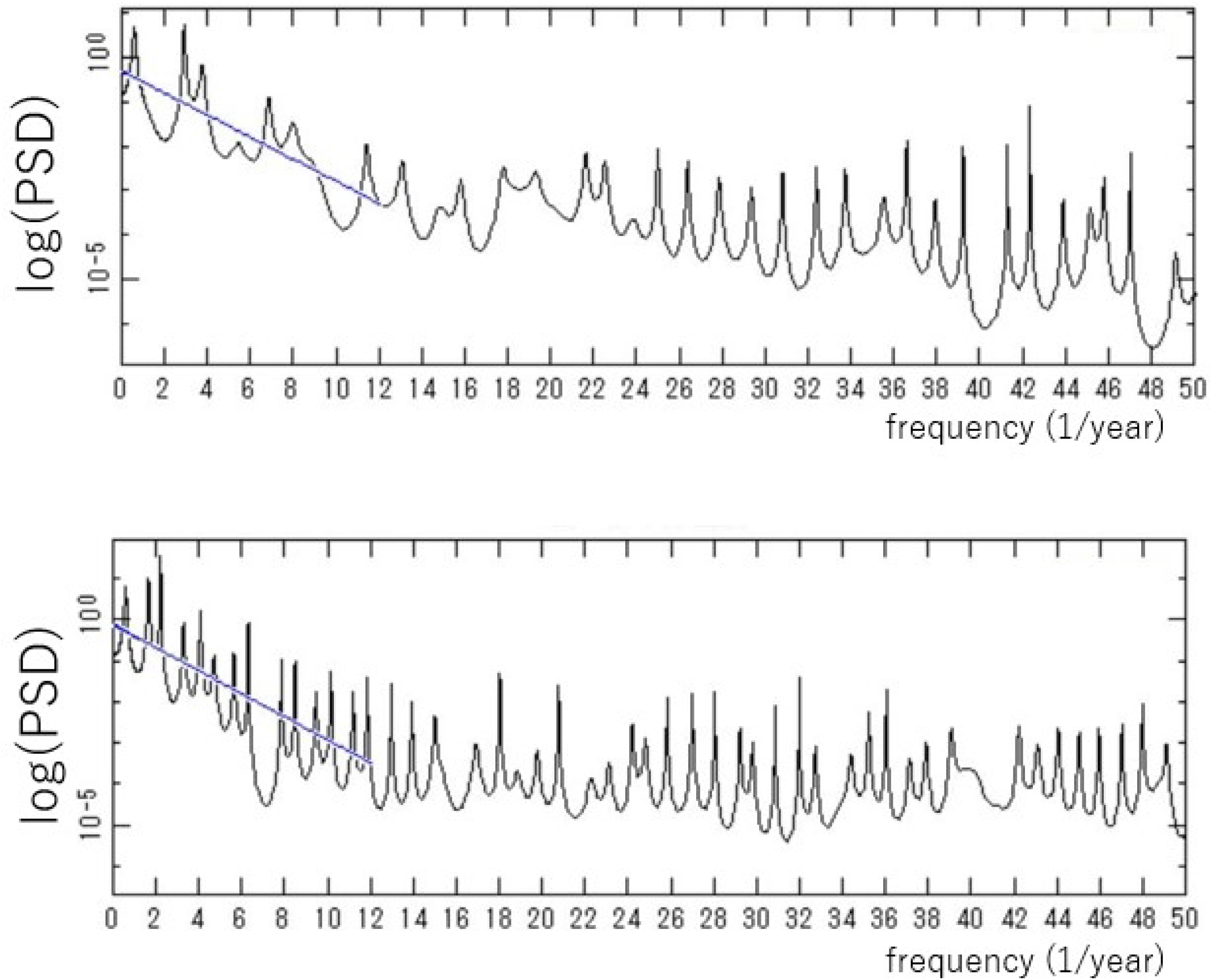
MEM-PSDs for two ranges of the residual data (*f* < 30.0). (a) Phase I (pre-Tokyo Olympic Games, 11 February 2020–22 July 2021). (b) Phase II (post-Tokyo Olympic Games, 23 July 2021-21 February 2023).

### Gradient of power spectral density

As seen in the PSDs for phases I and II in Figs 2a and 2b, respectively, the overall trend of each PSD indicates the exponential form

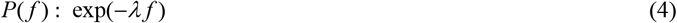

until the PSDs level off at the lowest limit determined by the accuracy of the present data, that is, the number of significant digits in the data. The values of *λ* for phases I and II are 0.25 and 0.28, respectively.

### Dominant spectral lines

Close-ups of the low-frequency regions of the PSDs in Figs 2a and 2b are shown in Figs 3a and 3b, respectively. For phase I (Fig 3a), the dominant spectral peak is observed at *f* =2.8, corresponding to a 0.36-year (4.3-month), with considerably large powers representing the amplitude of *x*(*t*) at each frequency. For phase II (Fig 3b), the dominant spectral peak is observed at *f* = 2.0, corresponding to a 0.50-year (6.0-month), with considerably large powers.

**Fig 3.**
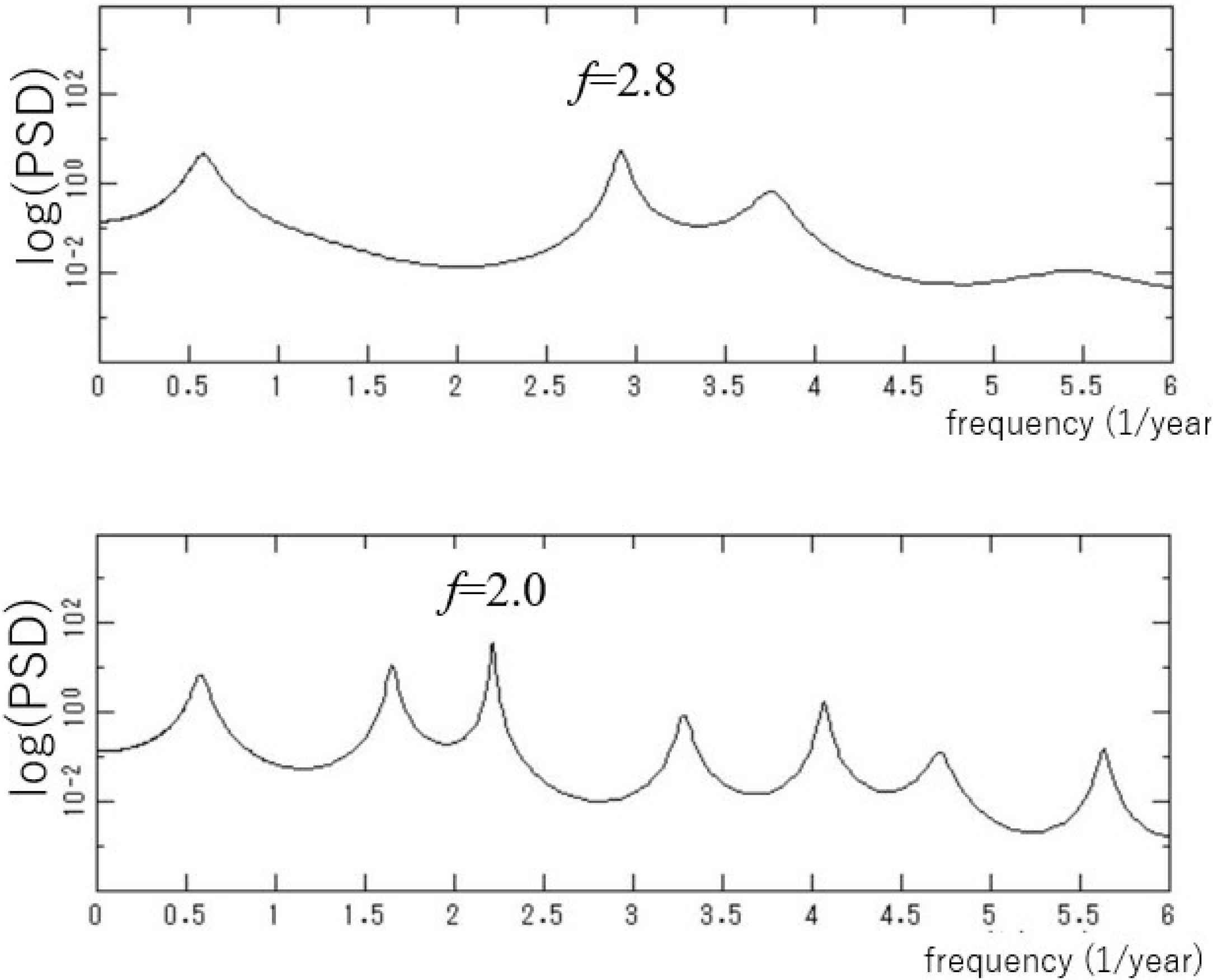
Close-up of the low-frequency region (*f* < 4.0) in Figs 2a and 2b: (a) Phase I (pre-Tokyo Olympic Games, 11 February 2020–22 July 2021). (b) Phase II (post-Tokyo Olympic Games, 23 July 2021–21 February 2023).

### Segment time series analysis

Periodic structures of the residual data were further investigated via segment time series analysis (Fig 1e). The residual data (Fig 1e) are divided into multiple segments, and the PSD is calculated for each segment. In this study, each segment represents the time interval of 365 days, and the starting point of two consecutive segments is delayed by six days. The PSDs thus obtained are shown as a 3D spectral array in Fig 4. In the 3D spectral array, the power is plotted against the frequency (horizontal axis) and time (right vertical axis). In the frequency range of 1.0 < *f* < 4.0, corresponding to the periods of 3.0 months to 1 year, spectral lines are clearly observed over the whole time range.

**Fig 4.**
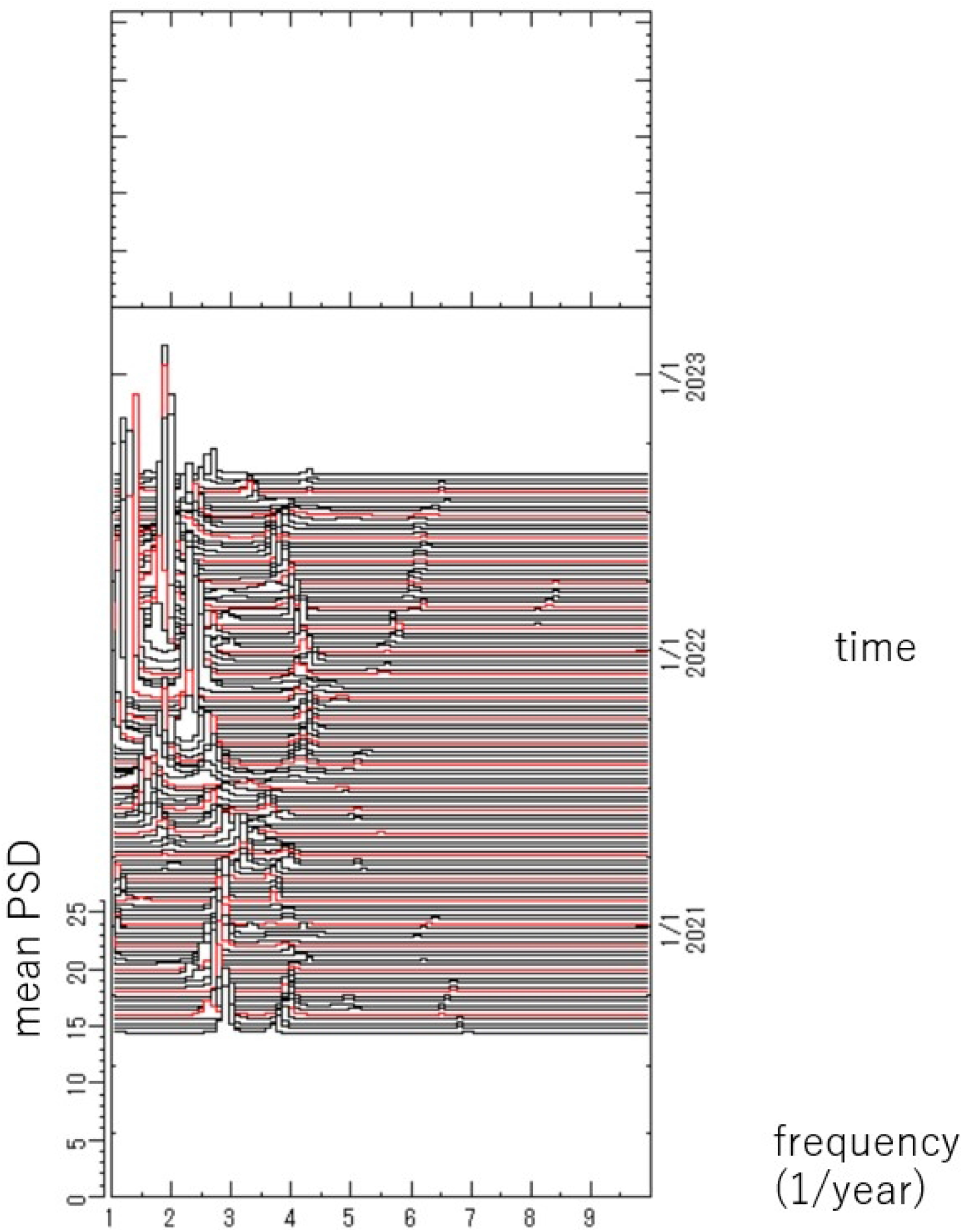
Three-dimensional spectral array for the residual data in the frequency range of 1.0 ≤ *f* ≤ 10.0.

The temporal variations of the frequency of dominant spectral lines observed in the 3D spectral array (Fig 4) are plotted in Fig 5. As seen in the figure, spectral peaks were observed around *f* = 3.0 (0.33 year) before May 2021. After May 2021, and beginning before the Tokyo Olympic Games started in July 2021, spectral peaks gradually migrated to the low-frequency range, and were observed to be around *f* = 2.2 from July to November 2021, and then remained relatively constant around *f* = 2.0 thereafter.

**Fig 5.**
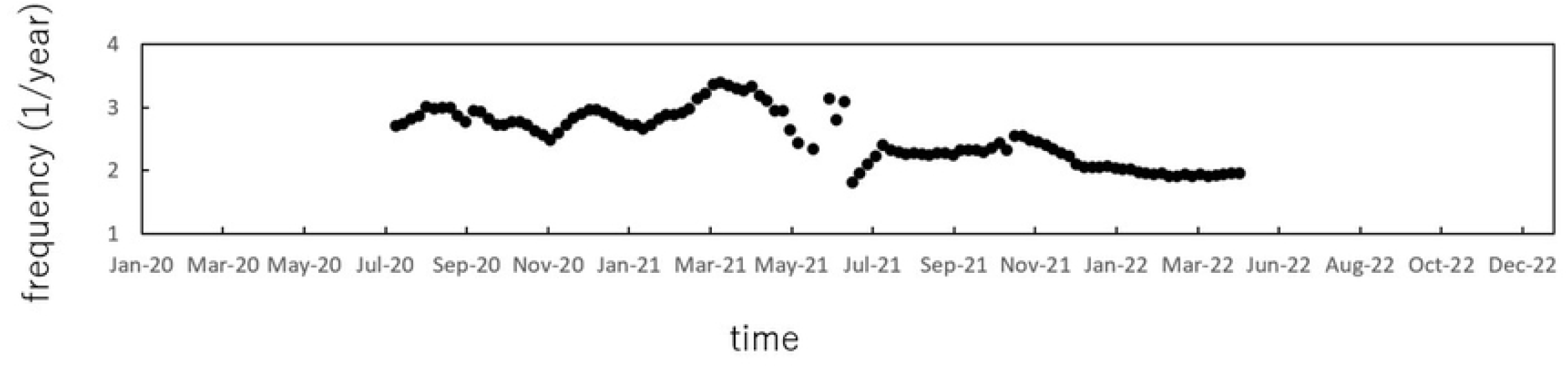
Temporal variations of the frequencies of dominant spectral peaks detected in the frequency range of 1.0 ≤ *f* ≤ 4.0.

## Discussion

The most notable result obtained in the present study is that, as observed in Fig 5, the gradual migration of the spectral line to the low-frequency range from 3.0 (0.33-year) to 2.0 (0.5-year) during May to July 2021 is already observable before the commencement of the Tokyo Olympic Games in July 2021 and immediately after the introduction of mass and workplace vaccination programs in April 2021, at a time when Japan’s vaccination rate was 4%. The vaccine rate increased quickly from May; the maximum number of daily inoculations was 1.6 million. By October 2021, more than 77 million people, equivalent to 61.8% of the targeted population, had completed the vaccination series [18]. The temporal behavior of periodic structures observed in Fig 5 indicates that theoretical studies for measles control programs, based on predictions that vaccination generates an increase in the inter-epidemic period (IEP), corresponding to the interval between major waves of an epidemic, may also apply to COVID-19 [19,20]. The IEP of measles epidemics has been investigated with time series analysis and mathematical models [21-27]. The IEP represents the amount of time required to accumulate a cohort of susceptible individuals that is sufficiently large to allow the measles virus to effectively spread over a community once it is introduced from outside. Our previous work investigated the IEP of measles epidemics in Japan and Wuhan in China using the present method of spectral analysis [19,20], and confirmed that the IEP increases as the vaccination ratio increases, as predicted by theoretical studies for a mathematical model of temporal variational structures of infectious diseases [21,22,28]. Based on the theoretical studies of measles and our preceding work in that disease, the present finding that period structures of the COVID-19 data of Japan changed temporally after May 2021 may be the effect of the increased vaccination rate in the previous month, April 2021.

With respect to the exponential characteristics of the PSDs for COVID-19 data (Figs 2a and 2b), our preceding work clarified that the PSDs for the time series generated from deterministic, nonlinear dynamical systems, such as the so-called susceptible/exposed/infectious/recovered (SEIR) epidemic model [29] and the Rössler, Lorenz and Duffing models [30,31], exhibit exponential characteristics. With respect to infectious disease epidemics, preceding research has confirmed that exponential spectral peaks are observed for incidence data of measles in Japan [15], Wuhan [19], New York City [29] and several communities in Denmark, the UK and the USA [32], as well as for chaotic and periodic time series generated by the SEIR epidemic model [29]. Thus, the present finding of exponential characteristics of the PSDs for COVID-19 (Figs 2a and 2b) suggests that the number of reported cases of COVID-19 in Japan is based on deterministic nonlinear dynamics.

For the magnitude of the PSD gradient λ, we clarified in our preceding work that λ decreases from the periodic state through the bifurcation process and reaches a minimum in the chaotic state, based on our preceding work on the SEIR model [29] and the Rössler model [30] throughout a series of period-doubling cascade through chaos. The decrease of the magnitude of λ can be considered to be the result of fluctuations mixed in a deterministic, nonlinear dynamical system [15]. With respect to the fluctuations, two possibilities have been postulated by the author’s group [15,30]; first, the amplitude fluctuation could be generated by an instability due to the nonlinearity of the system, as clarified in the result using the Rössler model, or the fluctuations could result from undeterministic noise. In both cases, the magnitude of λ decreases because the high-frequency components do not decline too rapidly [15,33]. In the present study, we confirmed that the magnitude of λ for the pre-Olympics period is smaller than that of the post-Olympics period. This result reflects the relatively high complex variations of the data in the pre-Olympics period, which appears to support Sapkota et al.’s finding that, among Japan’s 47 prefectures, the number of prefectures exhibiting chaotic characteristics was lower after the Tokyo Olympics than before [8]. A detailed study investigating chaotic characteristics for each prefecture in Japan is the subject of a forthcoming study.

## Conclusions

In general, biological phenomena are both nonstationary and nonlinear, and transit from one state to another in a complicated manner. Based on the results obtained in the present study, the periodic structures of epidemics of infectious diseases, including COVID-19, can be said to change over time. It is not appropriate to deal with the entirety of an overall time series incorporating such states. Thus, for elucidating a temporal evolution of nonlinear phenomena, it is preferable to deal with segments of time series of shorter data length via segment time series analysis, as performed in the present study. Investigation of temporal variational structures of disease epidemics with segment time series analysis can be expected to contribute to long-term and effective COVID-19 control programs in Japan.

In conclusion, the following three results are confirmed in the present study: First, the exponential characteristics of PSDs can be observed for the COVID-19 data of Japan in both pre- and post-Olympics periods, which is peculiar to the nonlinear dynamical process. Second, the magnitude of the gradient of exponential PSD for the pre-Olympics period is smaller than that of the post-Olympics period, because of the relatively high complex variations of the data in the pre-Olympics period caused by a deterministic, nonlinear dynamical system and/or undeterministic noise. Third, changes in the periodic structures of the COVID-19 data were already occurring before the Tokyo Olympic Games began in July 2021 and immediately after the mass and workplace vaccination programs were introduced in April 2021, indicating that the findings of theoretical studies for measles control programs may also apply to the COVID-19 data.

## Data Availability

The dataset of reported COVID-19 cases analyzed during the current study are contained in Supporting information files (S1 Dataset). The data are also available from ref. [12].

https://covid19.mhlw.go.jp/public/opendata/newly_confirmed_cases_daily.csv

